# Sublingual Ketamine for Depression and Anxiety: A Retrospective Study of Real-World Clinical Outcomes

**DOI:** 10.1101/2024.01.30.24301798

**Authors:** Lauren N. Swanson, Lila S. Jones, Jose Muñoz Aycart, Zhipeng Zhu, David M. Rabin, Taylor Kuhn

## Abstract

**Objective:** To evaluate the effectiveness of repeated at-home ketamine treatments for depression, generalized anxiety, and social anxiety and assess safety in terms of adverse effects and tendency towards long-term use.

**Methods:** This retrospective analysis included patients with depression, generalized anxiety, and/or social anxiety who received ketamine treatment (delivered at-home via low-dose, sublingual lozenges) through a private telehealth provider. Data was collected between May 2022 and April 2023. The primary outcome was change in depression, generalized anxiety, and social anxiety symptoms from baseline to three follow-up time points, measured via Patient Health Questionnaire (PHQ-9), Generalized Anxiety Disorder Assessment (GAD-7), and Social Anxiety Disorder Severity Scale (SAD-D-10), with analysis subgroups established based on baseline diagnosis. Secondary outcomes included side effects, adverse events, long-term use, well-being improvements, and comparison of outcomes between treatment-resistant and non-resistant depression cases.

**Results:** Of 431 patients (mean [SD] age, 43.6 [10.9] years; 49.2% women), 81 (18.8%) reported minor side effects resolving within 24 hours, and 397 concluded treatment in ≤ 6 months. Statistically significant improvement on the primary outcome was observed at all follow-ups in all three subgroups (p < 0.001). No significant differences were found between treatment-resistant and non-resistant depression outcomes.

**Conclusions:** Repeated sublingual ketamine significantly reduced depression, generalized anxiety, and social anxiety with no major adverse events and minimal tendency towards long-term use observed. These findings prompt further exploration of ketamine as an alternative or adjunct to medications such as SSRIs and benzodiazepines to minimize response delays and dependence risk.

## Introduction

Depressive and anxiety disorders are highly prevalent, debilitating conditions with considerable negative impacts on global disability and health-related quality of life.^1^ Severe depression and anxiety represent major risk factors for suicide, the second leading cause of death in Americans younger than 35.^2,3^

Despite the staggering burden of these conditions, treating them remains challenging.^4,5^ Selective serotonin reuptake inhibitors (SSRIs) are indicated as first-line medications, but lack widespread effectiveness, with roughly 40% of patients failing to meaningfully respond, and come with long delays before onset of therapeutic benefit and a range of undesirable side effects.^6–8^ Benzodiazepines are another class of medications used for managing anxiety disorders, and while they elicit a more rapid response than SSRIs, they are linked to a variety of adverse effects, including sedation/drowsiness/fatigue; emotional numbness; and impaired cognition, memory, and judgment.^9^ Long-term use carries high risk of physical dependence (i.e., tolerance and/or withdrawal) and high potential for abuse and addiction.^9^

Ketamine has been identified as a promising new treatment option that addresses some of the limitations of current medications, delivering rapid and robust antidepressant effects within hours following a single dose, notably in cases of treatment-resistant depression (TRD).^10^ Preliminary evidence suggests therapeutic benefits across diverse conditions, including generalized anxiety disorder (GAD) and social anxiety disorder (SAD).^11,12^

To date, the majority of studies have investigated ketamine delivered intravenously (IV), but less invasive routes, such as sublingual administration, are of interest to improve accessibility.^11^ The ability of sublingual ketamine to be self-administered allows for dosing in an at-home setting, which—assuming the presence of rigorous safety measures—could provide an attractive alternative to in-clinic infusions for select patients.^11^

While the efficacy and safety of ketamine have been well-established in depression, most research has focused primarily on its use in more severe, treatment-resistant cases,^13^ and clinical data on GAD and SAD is also limited.^14,15^

The aim of the present study was to examine clinical outcomes of a large group of patients with depression, generalized anxiety, and social anxiety treated with repeated, subanesthetic doses of sublingual ketamine in an at-home setting to assess effectiveness, safety, and feasibility.

## Methods

Data was obtained from a private telehealth practice that offered at-home ketamine treatment in the form of low-dose, sublingual lozenges for adults with depression, GAD, and SAD. All patients who were prescribed ketamine through the telehealth practice, Wondermed, between May 2022 and April 2023 were assessed for eligibility. All patients signed a detailed informed consent form granting the use of their personal health information for research purposes, subject to the requirements of applicable law. The study protocols were limited to secondary analysis of existing, de-identified patient data and involved no intervention or interaction with subjects. The institutional review board (IRB) of the University of California, Los Angeles, determined this study did not meet the definition of human subjects research, and as such, exemption from IRB review was granted.

### Participants

Patients seeking treatment through the practice underwent thorough evaluation by a licensed clinician to ensure the clinical appropriateness of at-home ketamine treatment. This included an eligibility questionnaire to screen for contraindications and an intake questionnaire that included symptom severity screens in preparation for a psychiatric consultation. Clinicians included physician assistants (PAs) and nurse practitioners (NPs) that underwent training by the telehealth practice on how to safely and effectively prescribe sublingual ketamine. Providers made the final determination as to whether the patient was well-suited for at-home ketamine treatment, and prescription issuance was limited to patients assessed as low-risk. Criteria used by Wondermed to determine eligibility for ketamine treatment are detailed in the eAppendix (Supplementary Material).

Study participants represent patients treated by the telehealth practice who completed at least one post-treatment assessment within 60 days of their baseline assessment.

### Analysis Subgroups

For analysis of primary outcomes, the sample was split into three subgroups according to the indication(s) for which they received treatment, defined by baseline scores that met or exceeded established thresholds for moderate disease severity (depression subgroup, PHQ-9 score ≥ 10; generalized anxiety subgroup, GAD-7 score ≥ 10; social anxiety subgroup, SAD-D-10 score ≥ 15).^16,17^ While most participants had comorbidities, resulting in substantial overlap between subgroups (**Table 1**), the rationale for further division was to prevent primary outcomes from being skewed by patients without comorbidities who presented with minimal symptoms of one or more disorders.

**Table 1.** Demographic and Clinical Characteristics of Study Sample.

| Characteristic | No. (%) patients (*n*=431) |
| --- | --- |
| **Biological sex** |  |
| Male | 219 (50.8) |
| Female | 212 (49.2) |
| **Age, years** |  |
| 20–29 years | 39 (9.1) |
| 30–39 years | 115 (26.7) |
| 40–49 years | 168 (39.0) |
| 50–59 years | 67 (15.5) |
| 60–69 years | 33 (7.7) |
| 70–79 years | 7 (1.6) |
| 80–89 years | 2 (0.5) |
| **Subgroup<sup>a</sup>** |  |
| Depression only | 22 (5.1) |
| Generalized anxiety only | 31 (7.2) |
| Social anxiety only | 29 (6.7) |
| Depression and generalized anxiety | 55 (12.8) |
| Depression and social anxiety | 25 (5.8) |
| Generalized and social anxiety | 38 (8.8) |
| All three | 147 (34.1) |

<sup>a</sup> Of the 431 patients included in the study, 249 met the criteria for inclusion in the depression subgroup (baseline PHQ-9 score $\geq 10$ ), 271 met the criteria for the generalized anxiety subgroup (baseline GAD-7 score $\geq 10$ ), and 237 met the criteria for inclusion in the social anxiety subgroup (baseline SAD-D-10 score $\geq 15$ ). Most (265 of 431; 61.5%) had comorbidities between 2 or three conditions based on baseline screening scores, resulting in overlap between the subgroups. A total of 84 patients did not meet the baseline score required for inclusion in any of the three subgroups and thus were excluded from primary outcome analysis and included in analysis of secondary outcomes only.

Patients in the depression subgroup were further classified into two cohorts based on treatment-refractory status to compare outcomes. The TRD cohort was defined as patients who reported prior use of two or more antidepressants without adequate response.^18^ While some TRD definitions include requirements related to dose and duration of previous trials, this data was not available in the present sample. The non-resistant cohort consisted of patients who: 1) reported inadequate response to only one previous antidepressant medication, 2) reported discontinuation of previous antidepressant(s) due to side effect intolerability, rather than lack of response, or 3) did not report previous antidepressant use.

The full flowchart of sample selection and subgroup designation is presented in **Figure 1**.

**Figure 1.**
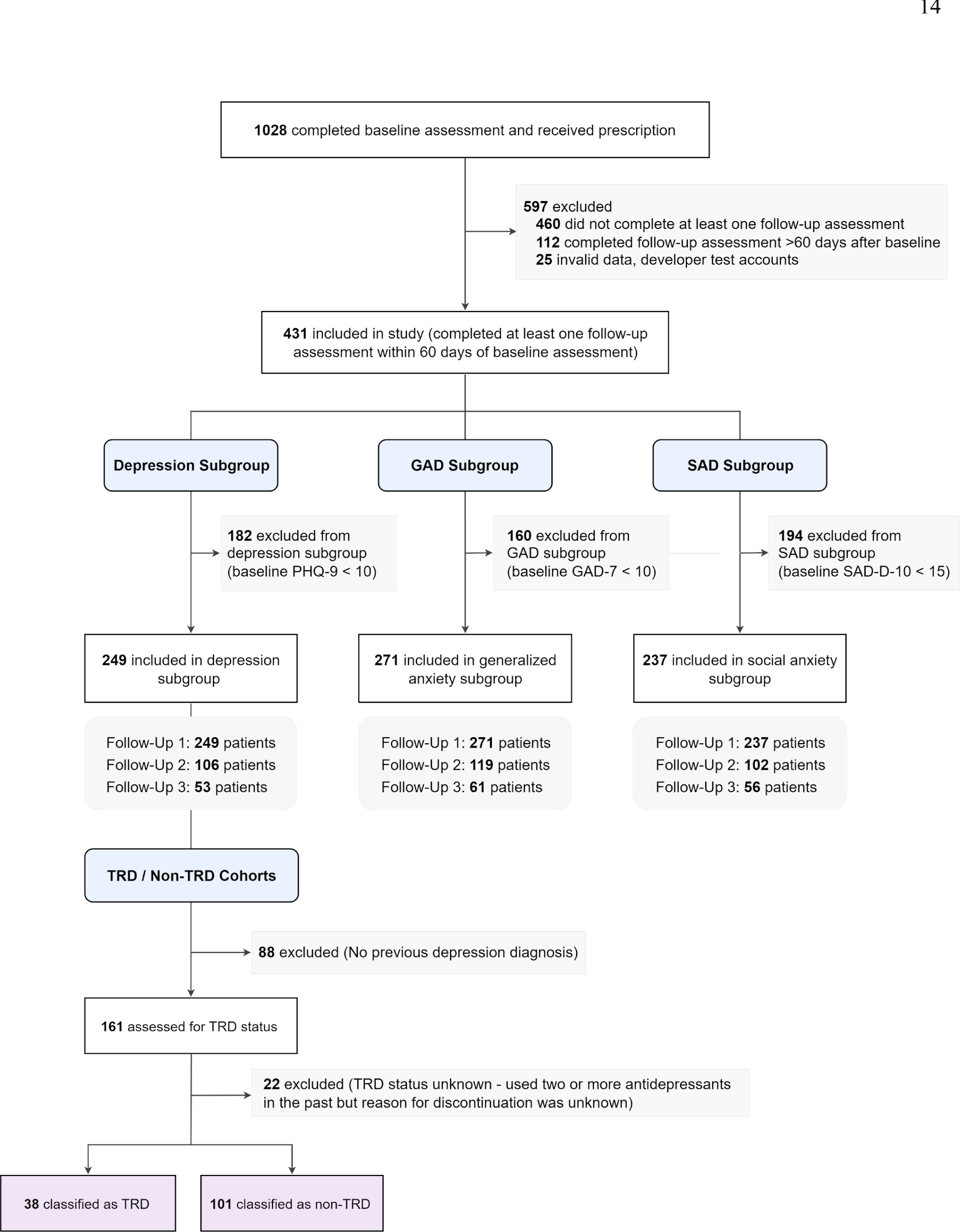
Flowchart of Participant Inclusion in Study Sample and Subgroup Analyses at Each Follow-Up Time Point Abbreviations: GAD, generalized anxiety disorder; Non-TRD, non-treatment-resistant depression; SAD, social anxiety disorder; TRD, treatment-resistant depression.

### Intervention

Patients determined to be good candidates for at-home treatment were prescribed a 1-month supply of ketamine. Most patients received the standard dose of 150–200 mg/lozenge at a once-weekly frequency, but dose could vary from 50–400mg at a maximum frequency of twice-weekly based on clinician discretion. Full data on medication prescription patterns is reported in Supplementary Table 1.

Patients wishing to renew their prescription for an additional month were required to complete a post-treatment assessment questionnaire and attend a remote follow-up consultation with their provider. Providers assessed the benefit of continued treatment, and dose was adjusted for follow-up prescriptions when appropriate. The intervention also included an integration program with daily exercises designed to help patients improve awareness of negative thought patterns and create new positive habits. Daily practices included journaling, meditation, breathwork, and therapy-based cognitive challenges. Hour-long soundscapes to listen to during treatment sessions were also provided.

### Data Collection and Outcomes

The study included all data collected from baseline, up to and including the first three follow-up assessments (occurring approximately 30, 60, and 90 days after baseline). The number of follow-ups included was limited to three due to patient numbers becoming much smaller thereafter (n < 50 in each subgroup). Demographic and clinical characteristics were collected at baseline.

The study’s primary outcome was change in depression, generalized anxiety, and social anxiety symptoms from baseline to three follow-up time points, measured using the 9-item Patient Health Questionnaire (PHQ-9), the 7-item Generalized Anxiety Disorder Assessment (GAD-7), and the 10-item Social Anxiety Disorder Severity Scale (SAD-D-10).

Secondary outcomes included safety, assessed using 1) data on side effects and adverse events (collected in a questionnaire administered after the first month of treatment, and at follow-up consultations), and 2) prevalence of long-term use (defined as ongoing treatment past 6 months, which was measured by number of prescriptions dispensed); as well as patient-reported changes across various well-being domains (collected at each follow-up). A final secondary outcome was comparison of treatment effects between treatment-resistant and non-resistant patients within the depression subgroup.

### Statistical Analyses

The data did not meet the assumptions required for the general linear model, thus necessitating nonparametric statistics for analysis of primary outcomes. To assess the statistical difference in scores at 1-, 2-, and 3-month follow-ups compared to baseline, the Skillings-Mack test was used, as it can account for missing data in analyses with repeated measurements.^19^ Dunn’s post hoc test was used to correct for multiple comparisons. Friedman’s ANOVA test was used for a subset of patients who completed three follow-ups (Supplementary Figure 1).

In addition to statistical significance, effect size (Cohen’s d, 95% confidence intervals) was also reported, along with other primary endpoints, including treatment response (50% or greater reduction in symptoms as reported on outcome measures from baseline to follow-up), remission (follow-up score less than 5), deterioration (score increase of 5 or greater), and meaningful clinical improvement (MCI), defined as a 20% or greater reduction in score compared to baseline (established in previous research for both PHQ-9 and GAD-7).^20,21^ While clinically meaningful change on the SAD-D-10 has not been established, prior work suggests this 20% threshold may be relevant across various psychiatric disorders.^21,22^

Descriptive analyses were conducted to determine the frequencies and distributions of sample population characteristics (e.g., age, sex, psychiatric disorder diagnoses); side effects and adverse events; and number of prescriptions dispensed. Changes in measures of well-being were also summarized using absolute frequencies and percentages of patients. For the treatment-resistant versus non-resistant depression analysis, a nonparametric independent t-test was used to compare PHQ-9 scores at each time point between the two cohorts.

All analyses were performed in R (Version 4.3.1), GraphPad Prism (Version 9.4.1), or Python 3, using the SciPy (1.11.2) and NumPy (1.23.5) libraries. A multiple comparison corrected *p-*value of < 0.05 was considered statistically significant. All statistical analyses are reported in figure legends and tables.

## Results

A total of 1,028 patient records were screened for eligibility. Of these, 597 patients were excluded from analysis due to: failure to complete at least one post-treatment assessment (n=460), completion of first post-treatment assessment > 60 days after completion of baseline assessment (n=112), and invalid data (n=25).

A total of 431 patients met criteria for inclusion into the final study sample (**Figure 1**), of which 212 (49.2%) were women, with mean [SD] age of 43.6 [10.9] years (**Table 1**). The mean (SD) time interval between each assessment for all participants was 33.9 (9.7) days.

Based on baseline severity scores, 249 patients met the criteria for inclusion in the depression subgroup, 271 met criteria for generalized anxiety, and 194 met criteria for social anxiety (**Figure 1**).

### Safety

Minor side effects were reported by 81 of 431 patients (18.8%), the most common being dizziness, headache, and nausea (Supplementary Table 2). One patient (0.02%) reported increased anxiety/depression, which was resolved within 24 hours of dosing. All side effects were resolved without reported medical assistance and did not persist for longer than 24 hours. No late-onset side effects were reported. There were no reports of misuse, development of addiction, or diversion, and no serious adverse events (suicidal ideation, suicides or attempted suicides, or self-harm behaviors) were observed. The majority of the sample (285 of 431) received ≤ 3 prescriptions before stopping treatment (66.1%), with 92.1% of patients (397 of 431) stopping treatment after ≤ 6 prescriptions.

### Primary Outcomes

In the depression subgroup, significant improvement in PHQ-9 score was found at all three follow-up time points compared to baseline (*p* < 0.001) (**Figure 2A**). Meaningful clinical improvement (MCI) was observed in 188 of 249 patients (75.5%) at 1 month, 85 of 106 patients (80.2%) at 2 months, and 47 of 53 patients (88.7%) at 3 months (**Table 2**).

**Table 2.** Clinical Outcomes for Each Subgroup at Three Follow-Up Time Points.

| Subgroup | *n*<sup>a</sup> | Mean (SE)<sup>b</sup> |  |  |  |  | No. (%) |  |  |  |
| --- | --- | --- | --- | --- | --- | --- | --- | --- | --- | --- |
|  |  | Baseline Score | Post-Test Score | Difference<sup>c</sup> | Cohen's *d* (95% CI) | *P* value<sup>d</sup> | MCI<sup>e</sup> | Response<sup>f</sup> | Remission<sup>g</sup> | Deteriorated<sup>h</sup> |
| Depression |  |  |  |  |  |  |  |  |  |  |
| 1 month | 249 | 16.02 (0.28) | 9.77 (0.29) | -6.26 (0.32) | 1.23 (1.07 to 1.40) | < .001* | 188 (75.5) | 99 (39.8) | 27 (10.8) | 2 (0.80) |
| 2 month | 106 | 15.93 (0.43) | 8.49 (0.42) | -7.44 (0.50) | 1.43 (1.16 to 1.71) | < .001* | 85 (80.2) | 49 (46.2) | 22 (20.8) | 0 (0.0) |
| 3 month | 53 | 16.08 (0.65) | 8.00 (0.61) | -8.08 (0.69) | 1.62 (1.20 to 2.03) | < .001* | 47 (88.7) | 26 (49.1) | 14 (26.4) | 1 (1.9) |
| Generalized Anxiety |  |  |  |  |  |  |  |  |  |  |
| 1 month | 271 | 14.17 (0.19) | 8.03 (0.24) | -6.13 (0.25) | 1.47 (1.30 to 1.64) | < .001* | 225 (83.0) | 118 (43.5) | 47 (17.3) | 1 (0.4) |
| 2 month | 119 | 14.10 (0.28) | 7.47 (0.33) | -6.63 (0.37) | 1.63 (1.36 to 1.90) | < .001* | 108 (90.8) | 60 (50.4) | 22 (18.5) | 2 (1.7) |
| 3 month | 61 | 14.28 (0.38) | 7.31 (0.57) | -6.97 (0.64) | 1.41 (1.05 to 1.76) | < .001* | 49 (80.3) | 39 (63.9) | 16 (26.2) | 1 (1.6) |
| Social Anxiety |  |  |  |  |  |  |  |  |  |  |
| 1 month | 237 | 24.00 (0.41) | 12.52 (0.48) | -11.48 (0.51) | 1.47 (1.28 to 1.65) | < .001* | 196 (82.7) | 125 (52.7) | 28 (11.8) | 4 (1.7) |
| 2 month | 102 | 24.12 (0.64) | 11.53 (0.72) | -12.59 (0.78) | 1.60 (1.31 to 1.90) | < .001* | 89 (87.3) | 63 (61.8) | 18 (17.65) | 2 (2.0) |
| 3 month | 56 | 24.41 (0.86) | 10.21 (1.00) | -14.2 (1.15) | 1.65 (1.25 to 2.06) | < .001* | 47 (83.9) | 37 (66.1) | 14 (25.0) | 0 (0.0) |

Abbreviations: CI, confidence interval; GAD, generalized anxiety disorder; MCI, meaningful clinical improvement; SAD, social anxiety disorder; SE, standard error.
<sup>a</sup>Number of patients.
<sup>b</sup>Values reflect scores on the PHQ-9 for the depression subgroup, on the GAD-7 for the generalized anxiety subgroup, and on the SAD-D-10 for the social anxiety subgroup.
<sup>c</sup>Difference refers to the mean change in scores from baseline.
<sup>d</sup>Adjusted *p* values with Bonferroni correction for multiple comparisons.
<sup>e</sup>MCI = meaningful clinical improvement, defined as a $\geq 20\%$ reduction in score compared to baseline.
<sup>f</sup>Response = treatment response, defined as a $\geq 50\%$ reduction in score compared to baseline.
<sup>g</sup>Remission is defined as a post-test score $< 5$ .
<sup>h</sup>Deterioration is defined as a worsening of $\geq 5$ points from baseline.
\* Statistically significant following Dunn post hoc test with Bonferroni correction for multiple comparisons.

**Figure 2.**
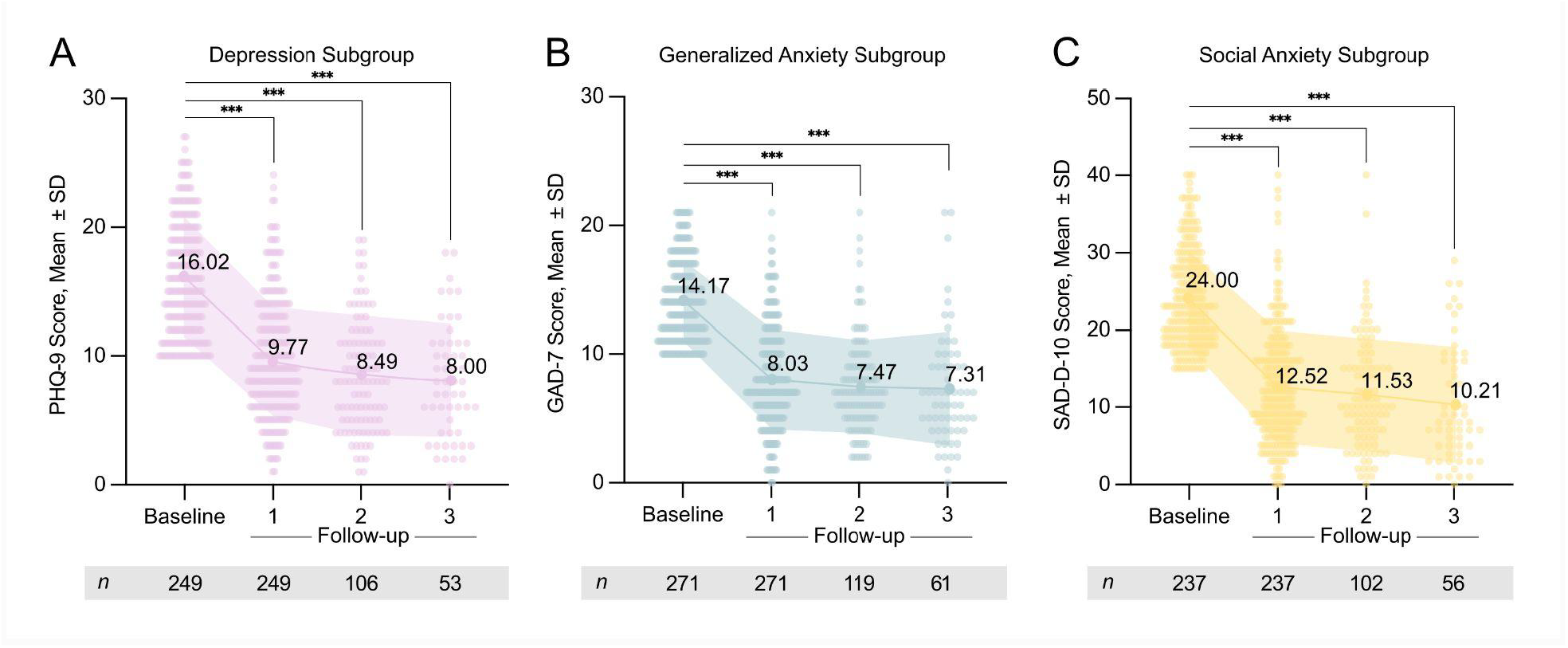
Changes in Depression, Generalized Anxiety, and Social Anxiety Symptom Severity for Each Subgroup for Patients Who Completed At Least One Post-Treatment Follow-Up Assessment A. PHQ-9 score at baseline, and at 1-, 2-, and 3-month follow-up in depression subgroup (Skillings-Mack test value=231.99, *n*=249, *p <* 0.0001). B. GAD-7 score at baseline and at 1-, 2-, and 3-month follow-up in generalized anxiety subgroup (Skillings-Mack test value=254.26, *n*=271, *p <* 0.0001). B. SAD-D-10 score at baseline and at 1-, 2-, and 3-month follow-up in social anxiety subgroup (Skillings-Mack test statistic=229.77, *n*=237, *p <* 0.0001). \*\*\**p* < 0.001, Dunn’s multiple comparison correction, Bonferroni adjusted. Abbreviations: SD, standard deviation.

The generalized anxiety subgroup also demonstrated statistically significant improvement from baseline at each follow-up (*p* < 0.001) (**Figure 2B**), with MCI achieved by 225 of 271 (83.0%), 108 of 119 (90.8%), and 39 of 49 (80.3%) of patients at months 1, 2, and 3, respectively (**Table 2**).

Statistically significant improvement across all three follow-ups was also observed within the social anxiety subgroup (**Figure 2C**). At 1 month, 196 of 237 patients (82.7%) achieved MCI, with 89 of 102 (87.3%) and 47 of 56 patients (83.9%) meeting criteria for MCI at months 2 and 3 (**Table 2**).

Across all subgroups, the highest rate of treatment response was observed at 3-month follow-up (**Table 2**). Patients meeting remission criteria increased at each follow-up time point, and deterioration was minimal (less than 3% of all patients) (**Table 2**), with no reports of patients or clinicians directly attributing deterioration to treatment. Several patients who deteriorated reported challenges in their personal lives or extra stress at work to their clinicians during follow-up appointments. Four patients who continued treatment after deteriorating demonstrated significant improvement at subsequent follow-ups, with 3 concluding treatment with mild symptoms and 1 achieving remission at final follow-up.

Data on the percentage of patients within each severity level category for each time point is presented in Supplementary Table 3.

### Improvements in Functioning and Well-Being

Of the full study sample (n=431), 90.7% (391 patients) self-reported positive change(s) in their overall well-being and functioning at 1 month, with higher numbers at months 2 (188 of 194, 96.9%) and 3 (100 of 101; 99.0%). Commonly reported changes included better ability to calm down when stressed (258 patients, 59.9%) and feeling more hopeful for the future (286 patients, 66.36%).

Full data on patient-reported positive changes is presented in **Table 3**.

**Table 3.** Improvements in Overall Well-Being and Psychosocial Functioning Reported by Patients at Three Follow-Up Time Points.

| Positive change | No. (%)<sup>a</sup> |  |  |
| --- | --- | --- | --- |
|  | 1 month<br>(n=431) | 2 month<br>(n=194) | 3 month<br>(n=101) |
| Any | 391 (90.7) | 188 (96.9) | 100 (99.0) |
| Feeling more hopeful for the future | 286 (66.4) | 137 (70.6) | 65 (64.4) |
| Better able to calm down when stressed | 258 (59.9) | 132 (68.0) | 71 (70.3) |
| Less irritable | 244 (56.6) | 105 (54.1) | 58 (57.4) |
| Experiencing less frequent / less severe mood swings | 209 (48.5) | 99 (51.1) | 62 (61.4) |
| Less reliant on or interested in alcohol or other drugs | 135 (31.3) | 70 (36.1) | 40 (39.6) |
| Better able to focus / concentrate | 121 (28.1) | 66 (34.0) | 38 (37.6) |
| More productive in work or studies | 118 (27.4) | 57 (29.4) | 36 (35.6) |
| Improved sleep | 104 (24.1) | 59 (30.4) | 35 (34.7) |
| More active / energetic | 96 (22.3) | 63 (32.5) | 37 (36.6) |
| Reduction in any chronic physical pain | 62 (14.4) | 31 (16.0) | 19 (18.8) |
| Other benefit(s)<sup>b</sup> | 44 (10.2) | 20 (10.3) | 8 (7.9) |

<sup>a</sup> Values in each column reflect the number of patients reporting each change at each follow-up time point.
<sup>b</sup> Examples of other benefits noted by patients included increased ability to process repressed emotions, feeling more gratitude in day-to-day life, less frequent social media use, reduced food cravings associated with emotional eating, and more consistently engaging in self-care.

### Depression Outcome Comparison Based on Treatment-Resistant Status

Of the 249 patients in the depression subgroup, 38 patients met criteria for TRD, and 101 met the criteria for the non-resistant cohort, with 110 patients excluded due to: (1) no previous depression diagnosis (n=88), (2) unknown TRD status (n=22) (**Figure 1**).

There was no significant difference in baseline depression severity between the treatment-resistant (mean [SD] PHQ-9 score, 17.03 [4.28]) and the non-resistant cohorts (mean [SD] PHQ-9 score, 16.68 [4.40]) (*p*=0.68), nor at any follow-up time points (TRD vs. non-TRD: 1 month, 9.18 [4.36] vs. 10.43 [4.58], [*p*=0.15]; 2 month, 9.15 [4.05] vs. 8.51 [4.20], [*p*=0.58]; 3 month, 8.67 [4.11] vs. 7.20 [3.98], [*p*=0.39]). These findings suggest comparable therapeutic effects regardless of treatment-resistant status.

Prevalence of long-term treatment was minimal in both cohorts, with just 3 of 101 non-resistant patients and 5 of 38 treatment-resistant patients continuing treatment past 6 months.

## Discussion

In the present study, repeated sublingual ketamine was associated with significant improvement in symptoms of depression, GAD, and SAD at three follow-up time points in an outpatient telehealth setting. Dosing sessions were well-tolerated, and no serious adverse events were reported within the study sample for the duration of the data collection period. Cases of significant symptomatic worsening were minimal. The rigorous eligibility criteria for treatment used by the telehealth provider may have contributed to these positive safety outcomes.

These findings suggest that with appropriate patient selection and monitoring, sublingual ketamine in an at-home setting can be a safe, practical, and clinically effective treatment option for patients with anxiety and depression. The clinical benefits observed in generalized and social anxiety symptoms are particularly noteworthy given the limited research on these indications.^15^ Also of note is the successful utilization of

PAs and NPs as the lead care providers for this intervention, demonstrating the potential for advanced practice practitioners like PAs and NPs to proficiently, effectively, and safely prescribe ketamine. In light of a worsening physician shortage, increased utilization of PAs and NPs to deliver ketamine treatment may represent an effective strategy to improve access to care and decrease costs.^23^ Future studies should investigate ketamine’s therapeutic potential across various psychiatric conditions in both open-label and randomized controlled trials.

While most consensus groups recommend consideration of ketamine only after other antidepressant options have been exhausted,^13,24,25^ our findings provide evidence that ketamine’s antidepressant efficacy is not exclusive to advanced cases of treatment-resistance. Earlier intervention with ketamine could be advantageous for select patients as a strategy to reduce the negative consequences of protracted ineffective treatment.^10^ For patients with suicidal ideation, ketamine’s rapid onset of effectiveness is especially crucial, reflecting a distinct advantage over the delayed efficacy of SSRIs.

At all three follow-ups, patients reported positive changes in functional outcomes and general well-being, including improved ability to regulate and manage emotions, increased productivity, and a more optimistic attitude about the future. Patients’ perceived benefits may have been mediated by the therapeutic integration resources included in the program (e.g., meditation, breathwork, and journaling exercises), as the value of integrating ketamine treatment with nonmedicinal therapeutic modalities and mindfulness-based practices has also been previously described.^26–28^ Further research should seek to clarify how to most effectively integrate ketamine with other supportive therapeutic practices to optimize treatment outcomes.

Previous studies found intermittent, repeated dosing regimens can prolong ketamine’s therapeutic effects; however, concerns exist about the safety and abuse potential of this approach, particularly in at-home treatment settings. In the present study, low inclination for continuous long-term treatment was observed, with 66.1% of patients concluding treatment within three months, and 92.1% within six months. Of note, as many as 83% of individuals prescribed a benzodiazepine for anxiety use the drug for longer than 6 months, despite clinical guidelines establishing treatment should not exceed 8–12 weeks.^29,30^ Given the high dependence and addiction potential of benzodiazepines, as well as the severe and potentially life-threatening effects associated with withdrawal,^9^ our findings provide preliminary evidence for the potential of ketamine as a safer alternative in specific contexts. We recommend that additional long-term studies be conducted to confirm the low abuse and misuse potential of repeated sublingual ketamine treatments in this patient population.

We acknowledge several limitations, including the retrospective observational design and the lack of a comparison group, resulting in within-subjects comparisons only. Generalizability is also limited by the characteristics of participants, all of whom actively sought at-home ketamine treatment through one private telehealth provider. Another notable limitation was that follow-up assessments were only administered by the telehealth practice when patients sought a prescription renewal. As a result, no post-treatment outcome data was available for patients who completed 1-month of treatment only, automatically excluding them from analysis. Individual reasons for treatment discontinuation could range from inadequate response, side effects, lack of perceived benefit, or symptomatic remission.

## Conclusion

This retrospective observational study evaluated the use of at-home sublingual ketamine in the context of real-world clinical practice, finding statistically significant benefits across depression, generalized anxiety, and social anxiety symptoms. Repeated dosing was well-tolerated, with no major adverse events reported. Future research is warranted to understand optimal dose, dosing frequency, duration of treatment, candidate selection, and the long-term outcomes and risks associated with treatment.

## Data Availability

All data produced in the present study are available upon reasonable request to the authors.

