## Supplementary Material for "Sublingual Ketamine for Depression and Anxiety: A Retrospective Study of Real-World Clinical Outcomes"

eAppendix. Inclusion and Exclusion Criteria for Ketamine Treatment Used by the Telehealth Practice

Supplementary Table 1. Prescription Doses and Dosing Frequencies

Supplementary Table 2. Side Effect Incidence and Time Until Resolved

Supplementary Table 3. Severity of Depression, Generalized Anxiety, and Social Anxiety Across All Participants at Baseline and Three Follow-Up Time Points

Supplementary Figure 1. Change in Depression, Generalized Anxiety, and Social Anxiety Symptom Severity in Patients Who Completed Three Post-Treatment Follow-Up Assessments

**eAppendix. Inclusion and Exclusion Criteria for Ketamine Treatment Used by the Telehealth Practice**

To be eligible for treatment, patients were required to meet the following inclusion criteria:

| |
|---|
| 18 years or older |
| Suffering from a depressive and/or anxiety disorder |

Exclusion criteria for ketamine treatment consisted of:

| |
|---|
| Pregnant, nursing, or trying to become pregnant |
| Interstitial cystitis |
| Uncontrolled hypertension |
| Raised intracranial or intraocular pressure (e.g., glaucoma) |
| Significant liver or kidney dysfunction |
| Respiratory depression from prior opioid overuse |
| Physical limitations due to heart disease, congestive heart failure, morbid obesity, asthma, chronic obstructive pulmonary disease (COPD) |
| Chronic use of narcotics |
| Primary psychotic disorder (schizophrenia, schizoaffective disorder) or borderline personality disorder |
| Experience of non-psychedelic-induced hallucinations or delusions |
| Bipolar disorder |
| National stressful events survey PTSD short scale (NSESSS) score $\geq 27$ |
| Past admission to the hospital for a psychiatric condition |
| Suicide attempt or thought of a plan to end life in the past year |
| History of dependence, addiction, or propensity for addiction |

### Supplementary Table 1. Prescription Doses and Dosing Frequencies

| Prescription[a] | First Rx *(n=431)* | Second Rx *(n=373)* | Third Rx *(n=234)* |
|---|---|---|---|
| **Dose / Lozenge** | | | |
| 50 mg | 1 (0.2) | 0 (0.0) | 0 (0.0) |
| 100 mg | 3 (0.7) | 0 (0.0) | 0 (0.0) |
| 125 mg | 2 (0.5) | 1 (0.3) | 0 (0.0) |
| 150 mg[a] | 138 (32.0) | 41 (11.0) | 15 (6.4) |
| 175 mg[a] | 39 (9.1) | 35 (9.4) | 18 (7.7) |
| 200 mg[a] | 128 (29.7) | 94 (25.2) | 37 (15.8) |
| 225 mg | 37 (8.6) | 46 (12.3) | 17 (7.3) |
| 250 mg | 48 (11.1) | 63 (16.9) | 54 (23.1) |
| 275 mg | 3 (0.7) | 13 (3.5) | 14 (6.0) |
| 300 mg | 20 (4.6) | 49 (13.1) | 37 (15.8) |
| 325 mg | 0 (0.0) | 3 (0.8) | 7 (3.0) |
| 350 mg | 3 (0.7) | 13 (3.5) | 13 (5.6) |
| 375 mg | 1 (0.2) | 0 (0.0) | 3 (1.3) |
| 400 mg | 8 (1.9) | 15 (4.0) | 19 (8.1) |
| **Dosing Frequency** | | | |
| Once weekly (4 lozenges total)[a] | 364 (84.5) | 265 (71.0) | 135 (57.7) |
| Every ~5 days (6 lozenges total) | 21 (4.9) | 73 (19.6) | 63 (26.9) |
| Twice weekly (8 lozenges total) | 46 (10.7) | 35 (9.4) | 36 (15.4) |

Abbreviation: Rx, prescription.

*Note:* Data represents the first three recorded prescriptions of all study participants. All values are expressed as No. (%), representing the number of participants prescribed each dosage.

[a] Standard starting dosage (150-200 mg/ lozenge, four lozenges total to be administered once weekly)

### Supplementary Table 2. Side Effect Incidence and Time Until Resolved

| Side Effects | No. (%)[a] | Time Until Resolved | | | |
|---|---|---|---|---|---|
| | | < 1 hr[b] | Several hrs[b] | < 1 d[b] | > 1 d[b] |
| Any | 81 (18.8) | - | - | - | - |
| Dizziness | 51 (11.8) | 22/51 (43) | 27/51 (53) | 2/51 (4) | 0/51 (0) |
| Nausea | 31 (7.2) | 12/31 (39) | 18/31 (58) | 1/31 (3) | 0/31 (0) |
| Headache | 23 (5.3) | 7/23 (30) | 11/23 (48) | 5/23 (21) | 0/23 (0) |
| Fatigue/ drowsiness | 4 (0.9) | 1/4 (25) | 2/4 (50) | 1/4 (25) | 0/4 (0) |
| Increased anxiety/ depression | 1 (0.2) | 0/1 (0) | 0/1 (0) | 1/1 (100) | 0/1 (0) |
| Light sensitivity | 1 (0.2) | 1/1 (100) | 0/1 (0) | 0/1 (0) | 0/1 (0) |

Abbreviations: d, day; hr, hour.

*Note:* All values are expressed as No. (%), indicating the number of participants who reported each event.

[a] Denominator used to derive percentages was the total number of participants in the study sample (n=431).

[b] Denominators used to derive percentages were the total number of patients who reported each specific side effect, based on row within table (dizziness [n=51], nausea [n=31], headache [n=23], fatigue/ drowsiness [n=4], increased anxiety/ depression [n=1], light sensitivity [n=1]).

#### Supplementary Table 3. Severity of Depression, Generalized Anxiety, and Social Anxiety Across All Participants at Baseline and Three Follow-Up Time Points

| Disease Severity | Baseline *(n=431)* | 1 month *(n=431)* | 2 month *(n=194)* | 3 month *(n=101)* |
|---|---|---|---|---|
| **Depression[a]** | | | | |
| None | 58 (13.5) | 112 (26.0) | 65 (33.5) | 40 (39.6) |
| Mild | 124 (28.8) | 190 (44.1) | 81 (41.8) | 37 (36.6) |
| Moderate | 106 (24.6) | 88 (20.4) | 38 (19.6) | 17 (16.8) |
| Moderately Severe | 82 (19.0) | 33 (7.7) | 10 (5.2) | 7 (6.9) |
| Severe | 61 (14.2) | 8 (1.9) | 0 (0.0) | 0 (0.0) |
| **Generalized Anxiety[b]** | | | | |
| None | 26 (6.0) | 113 (26.2) | 53 (27.3) | 38 (37.6) |
| Mild | 134 (31.1) | 213 (49.4) | 108 (55.7) | 46 (45.5) |
| Moderate | 157 (36.4) | 86 (20.0) | 27 (13.9) | 13 (12.9) |
| Severe | 114 (26.5) | 19 (4.4) | 6 (3.1) | 4 (4.0) |
| **Social Anxiety[c]** | | | | |
| None | 59 (13.7) | 121 (28.2) | 64 (33.2) | 43 (43.0) |
| Mild | 133 (30.9) | 212 (49.4) | 91 (47.2) | 38 (38.0) |
| Moderate | 142 (32.9) | 83 (19.3) | 33 (17.1) | 16 (16.0) |
| Severe | 81 (18.8) | 9 (2.1) | 3 (1.6) | 3 (3.0) |
| Extreme | 16 (3.7) | 4 (0.9) | 2 (1.0) | 0 (0.0) |

*Note:* All values are expressed as No. (%), representing the number of patients within each severity category (based on assessment scores) at each time point.  within each severity

[a] Based on PHQ-9, where scores of 0–4 are classified as none; 5–9 are classified as mild; 10–14 as moderate; 15–19 as moderately severe; and 20-27 as severe.

[b] Based on GAD-7, where scores of 0–4 are classified as none; 5–9 are classified as mild; 10–14 as moderate; and 15–21 as severe.

[c] Based on SAD-D-10, where scores of 0–4 are classified as none; 5–14 are classified as mild; 14–24 as moderate; 24–34 as severe; and 35–40 as extreme.

### Supplementary Figure 1. Change in Depression, Generalized Anxiety, and Social Anxiety Symptom Severity in Patients Who Completed Three Post-Treatment Follow-Up Assessments

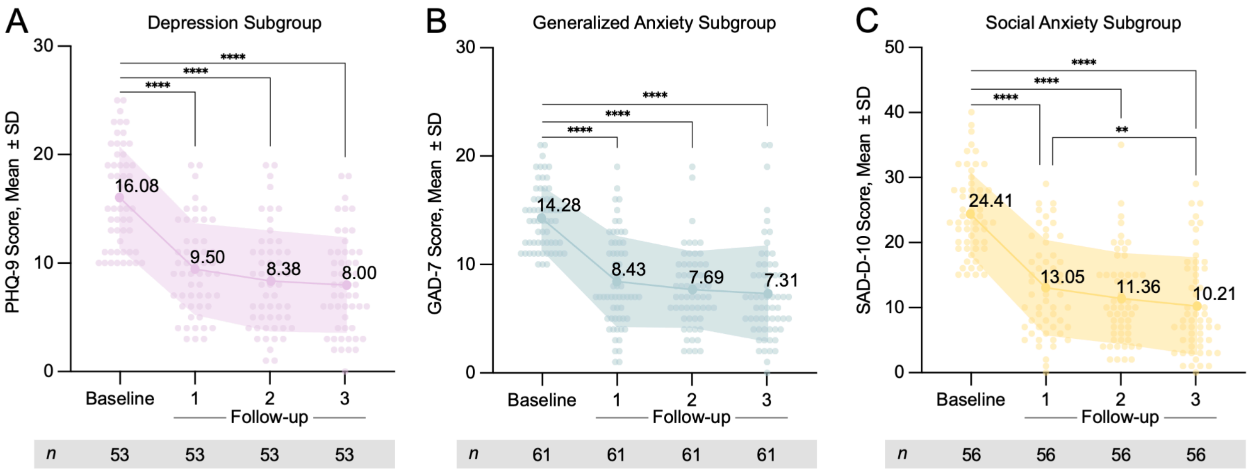


A. PHQ-9 score at baseline and at first, second, and third follow-ups in the depression subgroup (Friedman's test, $n$=53, $\chi 2$ = 83.60, $p < 0.0001$).

B. GAD-7 score at baseline and at first, second, and third follow-ups in the generalized anxiety subgroup (Friedman's test, $n$=61, $\chi 2$ = 80.10, $p < 0.0001$).

C. SAD-D-10 score at baseline and at first, second, and third follow-ups in the social anxiety subgroup (Friedman's test, $n$=56, $\chi 2$ = 84.02, $p < 0.0001$).

Dunn's multiple comparison correction, Bonferroni adjusted ****$p$ <0.0001, **$p$ <0.01.
